# Population anxiety and positive behaviour change during the COVID-19 epidemic: Cross-sectional surveys in Singapore, China and Italy

**DOI:** 10.1101/2020.04.14.20065862

**Authors:** Jane M Lim, Zaw Myo Tun, Vishakha Kumar, Sharon Esi Duoduwa Quaye, Vittoria Offeddu, Alex R Cook, May Oo Lwin, Shaohai Jiang, Clarence C Tam

## Abstract

**Background:** On 31 December 2019, an epidemic of pneumonia of unknown aetiology was first reported in the city of Wuhan, Hubei Province, People’s Republic of China. A rapidly progressing epidemic of COVID-19 ensued within China, with multiple exportations to other countries. We aimed to measure perceptions and responses towards COVID-19 in three countries to understand how population-level anxiety can be mitigated in the early phases of a pandemic.

**Methods:** Between February and March 2020, we conducted online surveys in Singapore, China and Italy with a total of 4,505 respondents to measure respondents’ knowledge, perceptions, anxiety and behaviours towards the COVID-19 epidemic, and identified factors associated with lower anxiety and more positive behavioural responses.

**Findings:** Respondents reported high awareness of COVID-19 and its accompanying symptoms, comparable information seeking habits and similarly high levels of information sufficiency, adherence to and acceptance of public health control measures. Higher self-efficacy was associated with lower anxiety levels in all three countries, while willingness to comply with restrictive measures and greater information sufficiency were associated with more positive behavioural changes to reduce spread of infection.

**Interpretation:** Population-level anxiety and behavioural responses to an outbreak can be influenced by information provided. This should be used to inform future outbreak preparedness plans, taking into account the importance of increasing population-level self-efficacy and information sufficiency to reduce anxiety and promote positive behavioural changes.

**Funding:** This was supported by the Saw Swee Hock School of Public Health, the Department of Communications and New Media, National University of Singapore, and the National Medical Research Council, Singapore

## Introduction

On 31 December 2019, an epidemic of pneumonia of unknown aetiology was first reported in the city of Wuhan, Hubei Province, People’s Republic of China.^1^ A rapidly progressing epidemic of what is now known as Coronavirus Disease 2019 (COVID-19) ensued within China, with multiple exportations to other countries. In response, a raft of control measures was implemented within China, including movement restrictions and mass quarantines of Wuhan and a number of other cities.

In Singapore, the first imported case, in a Chinese traveller from Wuhan, was reported on 23 January 2020, a week before COVID-19 was declared a Public Health Emergency of International Concern (PHEIC) by the World Health Organization (WHO).^2^ Clusters of local transmission within Singapore followed,^3^ with the first such cluster being identified on 5 February 2020. Increasingly stringent travel restrictions and mandatory social distancing for visitors and returning travellers have been gradually implemented as the epidemic has progressed, and extensive case detection, case isolation, contact tracing and quarantining have so far kept transmission in check and minimised community spread.

In contrast to Singapore, since February 2020 Italy has experienced an explosive epidemic with a death toll surpassing that of China.^4^ Mass quarantines and movement restrictions in the initially affected northern regions of Lombardy and Veneto have since been extended country-wide.

In addition to these epidemic control measures, clear and timely public communication from the public health authorities is a key feature of epidemic response. Particularly in the early phases of an epidemic, effective public messaging from trusted authorities is crucial for maintaining public assurance, but is challenging because of uncertainty in the level and sources of risk and the effectiveness of mitigation measures. Actions that reduce population anxiety and promote self-efficacy, however, could help to promote positive behaviour change to reduce the spread of infection. Between February and March 2020, we conducted online surveys in Singapore, China and Italy – three countries at different stages of the epidemic – to measure population knowledge, perceptions and behaviour in relation to the COVID-19 epidemic in each country.

## Methods

### Online panel surveys (Singapore and China)

Between 6 and 11 February 2020, we conducted an online survey in Singapore of public perceptions towards COVID-19 and epidemic control measures. The survey was disseminated to 1,766 members of an online survey panel maintained by the Singapore Population Health Studies (SPHS) unit of the Saw Swee Hock School of Public Health, National University of Singapore. Panel members were approached by email or SMS to complete a questionnaire administered on a secure web application, REDCap. The questionnaire was administered in English, took 10–15 minutes to complete and each respondent received a SGD5 (∼USD3.5) reimbursement for completing the survey. Data were anonymised before data analysis.

In China, a similar survey was administered between 10 February and 15 February 2020. The questionnaire was administered in Chinese to 3,492 respondents in the online survey panel maintained by a commercial research company (www.wjx.cn). This company sent emails including a link to the questionnaire to its online panel members. Respondents who completed the survey received a monetary incentive of 15 Chinese Yuan (∼USD2).

### Facebook surveys (Singapore and Italy)

Between 6 and 10 February 2020 and 14 and 18 February 2020, we also placed advertisements on the Facebook social media platform in Singapore. In Italy, we placed similar advertisements between 14 and 18 March 2020 for users in Lombardy and Veneto. The advertisements were displayed to Facebook users in the respective countries and contained a link to a survey questionnaire. No remuneration was provided for Facebook survey respondents.

### Survey questionnaire

The questionnaire comprised eight sections with questions on knowledge, sources of solicited and unsolicited information, self-efficacy and information needs, anxiety, confidence in authorities, acceptance of quarantine measures, behavioural changes in response to the epidemic, and socio-demographic characteristics. The knowledge section included questions regarding possible modes of SARS-CoV-2 transmission, personal protective measures and locally implemented public health control measures. For individuals who had actively sought information about the novel coronavirus, we asked what the main sources of information were and which they thought were the most trustworthy. We also asked about whether participants had received unsolicited information about the novel coronavirus, what type of information it was, and whether and why it was shared with others.

We assessed information needs by asking participants if they felt they had enough information regarding how to protect themselves and their families from coronavirus infection, what to do if they were infected, what public health measures had been put in place and why, and how confident they were that they could protect themselves from infection, as well as protect others if they themselves became infected. In addition, we included questions on superstition and fatalism in relation to health.

To measure anxiety, we adapted items from the State-Trait Anxiety Inventory^5^ to assess respondents’ level of worry in relation to the epidemic using a series of questions with a 5-point Likert response scale. To measure behavioural responses, we asked participants if they had modified or engaged in specific behaviours to reduce the risk of infection to themselves or others. These included personal hygiene behaviours and other protective measures, avoidance of workplaces, public spaces, social engagements or public transportation, changes to work-related or personal travel plans, and stockpiling of groceries, medications or personal protective equipment. Additionally, we included questions related to confidence in government and epidemic control measures, and acceptance of quarantine measures from previous published surveys.^6,7^

The basic questionnaire (Appendix A) was the same in all countries, but was modified to include country-specific response options or epidemic control measures. In China, it was not possible to include questions on confidence in government control measures. In addition, we omitted the majority of questions on knowledge from the Italian questionnaire as it was apparent by the time the survey was administered that the vast majority of the population would be aware of how COVID-19 was acquired, what the signs and symptoms were and what control measures had been implemented. The survey was administered in English in Singapore, in Chinese in China, and in Italian for respondents in Italy.

### Data analysis

We assessed representativeness of online survey and Facebook samples by comparing the sociodemographic characteristics of each survey sample with those from the respective national census in terms of age group, sex, educational level, and household income. In Singapore and China, ethnicity and housing type were additionally available. Since the majority of the Italian respondents were from Lombardia and Veneto, we compared these respondents to region-specific census data in Italy.

We assessed knowledge by tabulating variables related to possible modes of SARS-CoV-2 transmission, personal measures to reduce infection risk and control measures implemented by the local health authorities. We also tabulated the main sources of solicited (actively sought for) and unsolicited information about the novel coronavirus.

We calculated individual standardised scores for anxiety, confidence in authorities, acceptance of quarantine measures and behavioural responses by adding numerical responses within each of these sections. Higher scores corresponded to greater anxiety, greater confidence in authorities, greater acceptance of quarantine measures, and more positive behavioural changes in response to the epidemic respectively.

We performed a multivariable linear regression analysis to investigate whether anxiety was associated with risk perception, self-efficacy, confidence in authorities, acceptance of quarantine measures, superstition and fatalism, and use of different information sources, controlling for sociodemographic variables.

In a second multivariable linear regression, we assessed whether behaviour change in response to the epidemic was associated with anxiety, self-efficacy, confidence in authorities, acceptance of quarantine measures, superstition and fatalism, and use of different information sources, again controlling for sociodemographic variables.

Statistical analyses were performed in R version 3.6.1.^8^

### Ethics statement

Ethical approval for this study was provided by the Departmental Ethics Review Committee of the Saw Swee Hock School of Public Health, National University of Singapore (SPH-003; SPH-004), and the Departmental Ethics Review Committee of the Department of Communications and New Media, National University of Singapore (CNM-20200202-01).

## Results

We had a total of 4,505 eligible respondents in three countries: 2798 respondents from Singapore (1,529 from Facebook; 1276 from the online health panel), 1,089 from the China online health panel and 617 from Facebook in Italy. Response rates were based on the percentage of invited participants who completed the survey in the Singapore (71.8%) and China (31.2%) online panels, and the percentage of Facebook users who clicked on the survey link who subsequently completed the survey in the Singapore (22.4%) and Italy (9.8%) Facebook campaigns.

Compared to the respective countries’ census population **(Table 1)**, respondents in China were slightly younger, while Singaporean respondents were older. Across all three countries, survey samples had larger proportions of female respondents and respondents who reported having a tertiary education.

### Knowledge of COVID-19

Almost all respondents in Singapore and China were aware of COVID-19 and the implemented control measures. Most respondents identified common COVID-19 symptoms including cough (Singapore 95.5%; China 95.6%), fever (Singapore 95.5%; China 99.8%) and breathing difficulties (Singapore 88.3%; China 89.8%). Singaporean respondents were more likely to also identify milder symptoms such as sore throat (69.8%) and runny nose (74%) compared to their Chinese counterparts (28.2% and 16.6% respectively).

Respondents in both settings reported the following as common modes of COVID-19 transmission: coughing (Singapore 95%; China 91.2%), sneezing (Singapore 93.6%; China 97%), touching contaminated surfaces (Singapore 88.6%; China 86.8%) or close personal contact (Singapore 88.3%; China 84.8%). Places where Singaporean respondents thought people were most likely to contract COVID-19 were countries with outbreaks (89.3%), and public transport (75.8%). For Chinese respondents, public transportation (95.8%) and shopping malls (90.0%) were reported as places where infection risk was highest.

### Information needs/behaviours

More than three-quarters of all respondents (76.5% in Singapore; 96.1% in China; 94% in Italy) had actively searched for information about COVID-19. Most respondents named the internet (Singapore 64.4%; China 85.8%; Italy 71.2%) and social media (Singapore 52.4%; China 78.3%; Italy 59.5%) as their main information sources. However, when asked about trustworthiness, traditional media, including television, radio and print sources (Singapore 58.5.1%; China 72.2%; Italy 37.9%), and official government websites (Singapore 62.1%; China 53.9%; Italy 72.4%) were most often ranked as the most trustworthy sources of information.

Singaporean and Italian respondents reported receiving unsolicited information on COVID-19 most commonly through Facebook (Singapore 39.4%; Italy 50.2%) and WhatsApp (Singapore 40.2%; Italy 35.0%); Chinese respondents received unsolicited information via WeChat (87.1%) and Weibo (93.8%). Unsolicited information received included news stories (Singapore 72.2%; China 95.7%; Italy 64.0%), health advice (Singapore 69.7%; China 83.2%; Italy 58.4%) and advertisements for COVID-19 treatments (Singapore 28.9%; China 40.4%; Italy 35.5%). More than half of all the respondents said that they shared this unsolicited information with someone else.

### Self-efficacy/Information sufficiency

Most respondents felt that they had sufficient information about how SARS-CoV-2 is transmitted (Singapore 77.9%; China 88.4%; Italy 87.0%), what symptoms they might have if they were infected (Singapore 91.6%; China 88.2%; Italy 91.8%), what control measures health authorities were taking (Singapore 87.6%; China 76.6%; Italy 93.7%). However, a smaller percentage in China reported having sufficient information about the risk of infection (Singapore 84.3%; China 56.0%; Italy 86.7%) and why the authorities were taking specific control measures (Singapore 89.0%; China 61.2%; Italy 92.4%).

Self-efficacy among respondents was high; most felt that they had sufficient information about what to do if they thought they were infected (Singapore 86.7%; China 76.9%; Italy 93.1%), and how to protect themselves (Singapore 91.2%; China 97.9%; Italy 89.6%) and their family members (Singapore 86.1%; China 94.1%; Italy 90.1%) from COVID-19.

### Attitude and behaviour scores

Population anxiety towards COVID-19 was generally higher in China, while positive behavioural responses were higher in Italy. Respondents in Singapore had slightly higher scores for superstition and fatalism compared to Italy and China. In all three countries acceptance of restrictive public health measures and information sufficiency were similar, and confidence in authority was similar in Singapore and Italy. Individual scores for each country can be found in Table 1, and the distribution of scores in different domains can be found in **Figure 1**.

**Figure 1:**
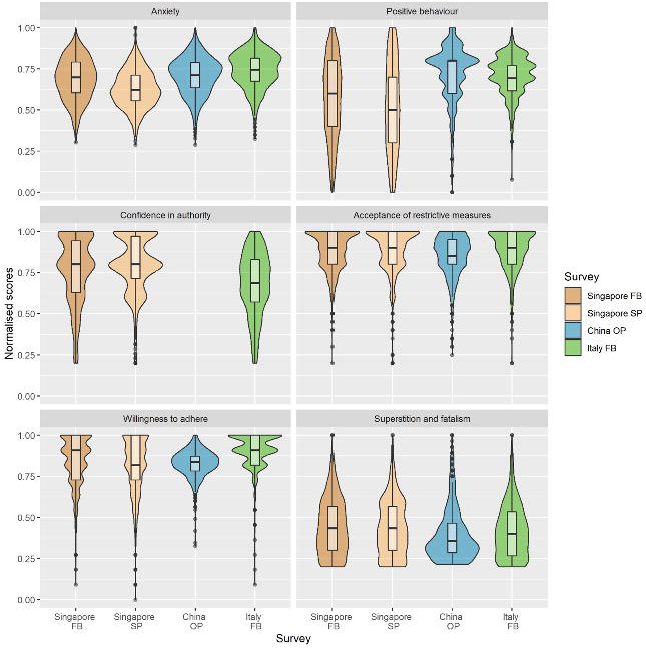
Distribution of respondents’ normalised scores for anxiety, positive behaviours, confidence in authority, acceptance of restrictive measures, willingness to adhere and superstition and fatalism.

### Factors associated with anxiety and positive behavioural response

Results of multivariable regressions on anxiety and positive behaviour scores are shown in **Tables 2 and 3**. Regression coefficients in both analyses represent the standard deviation change in mean score for the outcome variable per unit increase in corresponding dependent variables, adjusted for all other covariates in the model.

In all three countries, lower anxiety was associated with higher self-efficacy (confidence in the ability to protect oneself from infection) (Singapore Facebook: β=-0.30, 95% CI: -0.36 to -0.24; Singapore online panel: β=-0.28 (95% CI: -0.36 to -0.20); China: β=-0.24 (95% CI: -0.34 to -0.14); Italy: β= -0.17 (95% CI: -0.25 to -0.09)).

Higher anxiety was associated with higher scores for superstition and fatalism, and regarding traditional media as the most trustworthy information sources in both Singapore and Italy. Among Italian respondents, higher anxiety was seen particularly among those who considered messaging apps and friends and family the most trustworthy sources of information. For Singaporean Facebook respondents, higher confidence in authority was associated with lower anxiety scores, but the reverse was seen among Italian respondents **(Figure 2)**.

**Figure 2:**
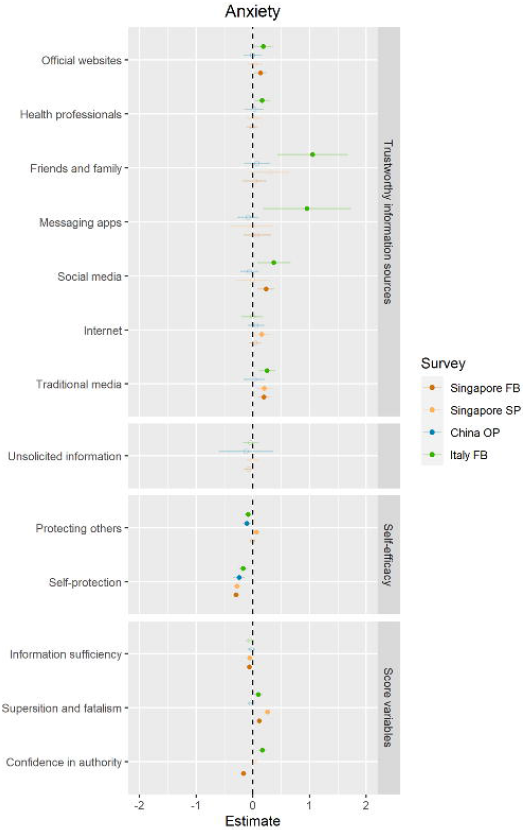
Forest plot for multivariable regression on anxiety.

Positive behavioural response was associated with willingness to adhere to control measures in all three countries **(Figure 3)**. In Singapore and Italy, greater self-efficacy (confidence in the ability to protect family members if respondents became infected), greater acceptance of restrictive public health measures, and higher anxiety were also associated with more positive behavioural responses. In China, information sufficiency was the factor most strongly associated with positive behaviour. Conversely, superstition and fatalism were negatively associated with behavioural response in China and Italy, while in Singapore respondents displaying greater confidence in authority had less positive behavioural responses. We did not observe strong associations between information sources and positive behavioural response, except in Singapore, where reliance on traditional and social media as trustworthy information sources was associated with more positive behaviour.

**Figure 3:**
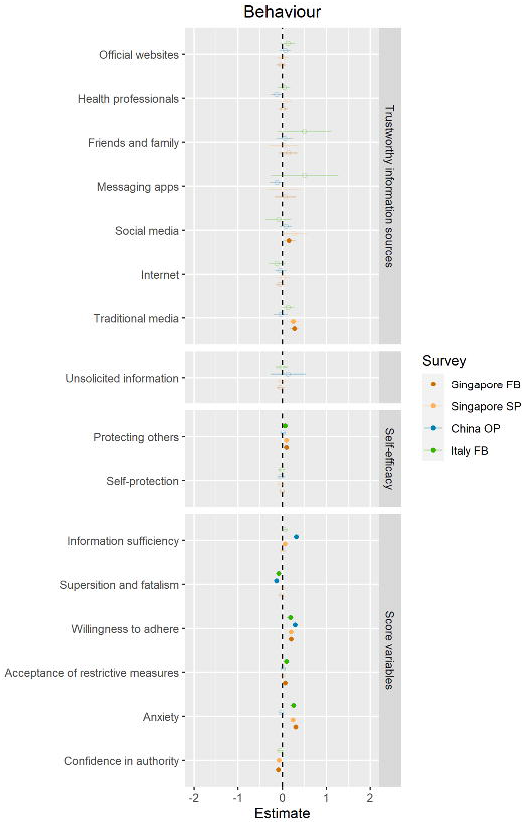
Forest plot for multivariable regression on positive behavioural response.

## Discussion

We assessed the knowledge, attitudes and overall perceptions of the general public in Singapore, China and Italy during the initial months of the COVID-19 epidemic. Results showed that respondents across all three countries had comparable information seeking habits and similarly high levels of information sufficiency, adherence to and acceptance of public health control measures. However, there were some key differences in their awareness about COVID-19 symptoms – although most Singaporean and Chinese respondents recognised fever, cough and breathing difficulties as COVID-19 symptoms, Chinese respondents were less likely to report runny nose and sore throats as symptoms of COVID-19, perhaps reflecting a greater emphasis on severe disease manifestations in China.

Current evidence indicates that SARS-CoV-2 transmission risk is most likely to result from close contact in households, large social gatherings or workplaces.^3^ However, respondents in Singapore and China reported that transmission was most likely in places such as shopping malls and public transportation and least likely at home. This corresponds with respondents’ self-reported mask wearing intentions; most respondents in both countries said that they would not wear a face mask at home to reduce their risk of infection but would wear them out in public, signaling a need for better messaging around infection prevention within the household.

Our findings indicate that having sufficient information about what control measures are in place and why, and individual beliefs in the ability to protect oneself and others are strongly associated with lower anxiety during an epidemic across all three settings. This is likely to be dependent on the availability of trustworthy information. Respondents in our surveys obtained information from a range of sources, including the internet, television, radio and print, social media and peers. Despite this, official government sources were considered by the majority to be the most trustworthy sources of information.

Greater self-efficacy was associated with reduced anxiety, while higher superstition and fatalism scores were associated with higher levels of anxiety in Singapore and Italy. Previous studies show that fear and anxiety during a pandemic can be mitigated by self-efficacy^6^ and information sufficiency, while tendencies towards fatalism reduce self-efficacy and increase anxiety.

In all three countries, higher acceptance of restrictive control measures and information sufficiency were strongly associated with greater positive behavioural response. Respondents’ anxiety was also a catalyst in their adoption of positive behavioural responses to reduce risk of infection – in particular, Singaporean respondents with higher levels of anxiety and self-efficacy were more likely to adopt positive preventive measures, consistent with previous studies done during the 2009 influenza A/H1N1 pandemic.^9^ Conversely, Singaporean respondents with greater confidence in authority were less likely to adopt positive behaviours; high trust in government control measures could lead to passivity in individual response through behavioural change. In both Chinese and Italian respondents, higher scores of superstition and fatalism were also associated with less positive behavioural change. This agrees with previous literature suggesting that belief that health status is largely determined by external forces makes individuals less likely to adopt behaviours that can positively impact one’s health outcomes.^10^

Overall anxiety levels and adoption of positive behaviours were strongly influenced by information sufficiency in all three countries, showing that population-level risk perception, self-efficacy and response to an outbreak can be intensified or attenuated by the quantity and quality of information provided.^11,12^ These findings highlight the importance of disseminating authoritative information from trusted health and government authorities through a variety of online and traditional media outlets, especially during the initial phase of an outbreak. Clear information about signs and symptoms of the disease, risk reduction measures, protective behaviours and why specific control measures are being taken should be provided via these channels.

While concerns have been raised about the potential for social media and messaging platforms to disseminate misinformation, we did not find strong or consistent evidence in our surveys that information obtained through these platforms increased anxiety or resulted in less positive behavioural responses. This could reflect a bias in the sample of participants who chose to respond to the survey, but could also indicate the fact that these platforms can have both positive and negative influences. While many respondents used these platforms for information, they were not generally regarded as the most trustworthy information sources, suggesting that individuals may not necessarily act solely on information received through social media. However, a large fraction of respondents reported receiving unsolicited information through these platforms. We were unable to capture details on the content of this information, so cannot make conclusions about its accuracy. Despite this, a concerning number of respondents reported that they received advertisements about purported treatments for COVID-19.

The use of online survey methodologies has specific advantages during an epidemic, when there is a need to collect information quickly but when there may be challenges to the safe conduct of in-person community surveys. Our internet-based sampling strategy and survey dissemination via an online panel to a large number of respondents enabled the rapid deployment of a survey to track responses in near real-time, allowing us to study risk perception and behavioural changes during the early phases of an outbreak. A limitation of this approach is the trade-off in terms of population representativeness. Our sample shows a pronounced over-representation of respondents with at least a university education in both Singapore and China, which may affect the generalisability of our findings.

## Conclusion

This study presents important information about risk perceptions and behavioural responses during the initial phases of an epidemic of a novel virus across different settings. Results from this study should be used to inform future outbreak preparedness plans. In particular, policymakers should take into account the importance of increasing population-level self-efficacy and information sufficiency to mediate anxiety and promote positive behavioural changes.

## Data Availability

All relevant data are within the manuscript and its Supporting Information files.

## Acknowledgements

We thank the SPHS unit for their assistance in pre-testing and data management for the questionnaire fielded to the online panel.

## Authors’ contributions

All authors attest they meet the ICMJE criteria for authorship. All authors contributed to the design and implementation of the research. ZMT and CCT developed and conducted the analysis, JL and CCT drafted the manuscript. All authors provided critical feedback on the manuscript.

## Figure and table legend

**Table 1:** Comparison of sample demographic characteristics to the respective countries’ census population

**Table 2:** Multivariable regression results for anxiety

**Table 3:** Multivariable regression results for positive behavioural response

## Appendix A

Questionnaire

## Supplementary online material

Sample demographic characteristics and scores

## References

1 Wu JT, Leung K, Leung GM. Nowcasting and forecasting the potential domestic and international spread of the 2019-nCoV outbreak originating in Wuhan, China: a modelling study. The Lancet 2020; 395: 689–97.

2 Wong JE, Leo YS, Tan CC. COVID-19 in Singapore—current experience: critical global issues that require attention and action. Jama 2020.

3 Pung R, Chiew CJ, Young BE, et al. Investigation of three clusters of COVID-19 in Singapore: implications for surveillance and response measures. The Lancet 2020.

4 Remuzzi A, Remuzzi G. COVID-19 and Italy: what next? The Lancet 2020.

5 Spielberger CD, Sydeman SJ, Owen AE, Marsh BJ. Measuring anxiety and anger with the State-Trait Anxiety Inventory (STAI) and the State-Trait Anger Expression Inventory (STAXI). Lawrence Erlbaum Associates Publishers, 1999.

6 Bults M, Beaujean DJ, de Zwart O, et al. Perceived risk, anxiety, and behavioural responses of the general public during the early phase of the Influenza A (H1N1) pandemic in the Netherlands: results of three consecutive online surveys. BMC public health 2011; 11: 2.

7 Tracy CS, Rea E, Upshur RE. Public perceptions of quarantine: community-based telephone survey following an infectious disease outbreak. BMC Public Health 2009; 9: 470.

8 RStudio Team. RStudio: Integrated Development for R. Boston, MA: RStudio, Inc., 2015 http://www.rstudio.com/.

9 Jones JH, Salathe M. Early assessment of anxiety and behavioral response to novel swine-origin influenza A (H1N1). PLoS one 2009; 4.

10 Keeley B, Wright L, Condit CM. Functions of health fatalism: fatalistic talk as face saving, uncertainty management, stress relief and sense making. Sociology of health & illness 2009; 31: 734–47.

11 Griffin RJ, Neuwirth K, Dunwoody S, Giese J. Information sufficiency and risk communication. Media psychology 2004; 6: 23–61.

12 Choi D-H, Yoo W, Noh G-Y, Park K. The impact of social media on risk perceptions during the MERS outbreak in South Korea. Computers in Human Behavior 2017; 72: 422–31.

